# Covid-19 Vaccine Booster Cadence by Immunocompromised Status

**DOI:** 10.1101/2023.04.18.23288615

**Authors:** Matan Yechezkel, Jeremy Samuel Faust, Doron Netzer, Talish Razi-Benita, Erez Shmueli, Ronen Arbel, Dan Yamin

**Affiliations:** Department of Industrial Engineering, Tel Aviv University, Tel Aviv, Israel; Brigham and Women’s Hospital, Division of Health Policy and Public Health, Department of Emergency Medicine, Harvard Medical School, Boston, Massachusetts; Community Medical Services Division, Clalit Health Services, Tel Aviv, Israel; MIT Media Lab, Cambridge, MA, USA; Maximizing Health Outcomes Research Lab, Sapir College, Sderot, Israel; Center for Combatting Pandemics, Tel Aviv University, Tel Aviv, Israel

## Abstract

**Background:** Data suggest that vaccine effectiveness against Covid-19-associated hospital admission and mortality is augmented with booster doses, but the benefit wanes within several months. However, the CDC recently concluded that second doses of bivalent vaccines this Spring were not warranted because existing data were insufficient to analyze the benefits of such a strategy. Therefore, our objective was to assess whether routinely boosting high-risk populations at least every 6 months may be warranted, depending on age and immune status.

**Methods:** Utilizing a database of 3,574,243 members of Clalit Health Services (CHS), we analyzed the medical records of individuals who received none, or at least one dose of the BNT162b2 mRNA COVID-19 vaccine between January 1, 2021, and April 5, 2022. We examined the risk of moderate-to-severe Covid-19 hospitalization or death stratified by age group, immune status and time since receipt of the last vaccine dose during the early Omicron wave in Israel (December 20, 2021 to April 5, 2022). The number needed to vaccinate (NNV) was calculated as the inverse of the absolute risk reduction for various subgroups and Covid-19 waves.

**Results:** Eligibility criteria were met by 3,381,480 CHS members. The absolute risk of Covid-19 moderate-to severe hospitalization or death during the Omicron wave increased with age, immunocompromised status, and time since receipt of the last vaccine dose. The NNVs varied greatly by age and immune status and were contingent on various disease prevalence scenarios. Among the severely immunocompromised, boosting at the start of the Omicron wave had an NNV ranging from 87 (95% CI 70-109) in persons ages ≥80 to 1,037 (95% CI 999 -1,513) in persons ages 12-59. In the lower prevalence periods, the NNV for 6-month booster cadencing remained favorable for immunocompromised people in all age groups and immunocompetent people ages ≥60.

**Conclusions:** Our study provides evidence for the potential benefit of a routine 6-month cadence for Covid-19 boosters for the highest-risk groups, and possibly more frequently, even during relatively lower Covid-19 prevalence.

## BACKGROUND

Data suggest that vaccine effectiveness against Covid-19-associated hospital admission is augmented with booster doses but that the benefit wanes within several months, including among the immunocompromised and older groups.^1–3^ Because of baseline risk differences, however, this waning may leave immunocompromised and older persons at unacceptably high-risk several months after booster receipt compared to immunocompetent persons, especially given the loss of effectiveness offered by monovalent antibodies authorized for that population.^4^ Accordingly, the US Food and Drug Administration (FDA), the US Centers for Disease Control and Prevention (CDC), and many global peer nations authorized additional boosters for immunocompromised persons during the 2021-mid-2022 period.^5,6^

Since September 2022, bivalent Covid-19 boosters have been recommended for all persons age 5 and older in the United States regardless of age, risk, or immune status.^7,8^ The CDC recently concluded that second doses of bivalent vaccines this spring were not warranted for any age or risk group, since the existing data were insufficient to analyze the benefits of such a strategy.

Therefore, our objective was to assess whether Covid-19 boosters are currently warranted and whether routinely boosting high-risk populations at least every 6 months may be warranted, depending on age and immune status.^10^

## METHODS

### Study Population and Data sources

We analyzed the medical records of individuals aged 12 years or older who are members of the largest healthcare organization in Israel, Clalit Health Services (CHS). CHS is the largest healthcare provider in Israel, serving about 52% of the population (approx. 4.7 million members). CHS’s members are representative of the Israeli population and reflect all demographic, ethnic, and socioeconomic groups and levels. Records are automatically collected and updated monthly in the databases of all CHS medical facilities across the country.

The data are coded, pseudonymized, viewed, stored, and processed within the CHS research room. CHS uses the International Classification of Diseases, Ninth Revision (ICD-9), and Clinical Modification. Procedures are coded using Current Procedural Terminology codes. We accessed each patient’s demographic information and information on potential chronic illnesses. Covid-19 data (vaccinations, infections, hospitalizations, and death) is collected centrally by the Israeli Ministry of Health and updated in the CHS database daily.

As the retrospective data was pseudonymized, the institutional Helsinki review board and data utilization committee approved the use of the retrospective cohort data without requiring specific consent from the members of Clalit Health Services (protocol number O139-21-CHS).

### Definition of immunocompromised status

We classified individuals based on their immunocompromised statuses: ‘severely immunocompromised’, ‘mild or moderately immunocompromised’ and immunocompetent. The definition of ‘severely immunocompromised’ is based on the US CDC’s and the Advisory Committee on Immunization Practices’ (ACIP) definition.^7,11^ Specifically, severely immunocompromised individuals include people with an active malignancy, receipt of a solid organ transplant, individuals diagnosed with hematopoietic malignancies (i.e., chronic lymphocytic leukemia, non-Hodgkin lymphoma, multiple myeloma, acute leukemia), or people diagnosed with cystic fibrosis or sickle cell anemia. We defined ‘mild or moderately immunocompromised’ as individuals with at least one of the following conditions: Rheumatoid Arthritis, Ulcerative Colitis, Crohn’s Disease, Celiac Disease, and Systemic Lupus Erythematosus. This definition is based on the US CDC’s and the Advisory Committee on Immunization Practices’ (ACIP) definition.^7,11^ We classify individuals as immunocompetent if they are not immunocompromised (severely or not severely).

### Eligibility Criteria

To be included in our analysis, individuals needed to be active CHS members during the entire study period, except for death. Additionally, individuals had to receive none, or at least one dose of the BNT162b2 mRNA Covid-19 vaccine between January 1, 2021, and April 5, 2022. We excluded individuals who received any other Covid-19 vaccine dose than the BNT162b2 mRNA COVID-19 vaccine. Additionally, we excluded individuals with mild or moderate immunocompromise statuses from the primary analysis. A secondary analysis including all person with immunocompromise (mild, moderate, or severe) was conducted.

### Definition of Waves

For the primary analysis, we define the ‘early Omicron wave’ as the period between December 20, 2021 and April 5, 2022 (BA. I and BA.2 predominance period.^12^ The ‘low prevalence’ Covid-19 period is defined as the time From September 1, 2022, to December 17, 2022 (which maintained the same period length as the early Omicron wave; BA.5 and BQ.1 predominance period).^12^ Both definitions were based on COVID-19 prevalence data published by Israeli Ministry of Health.^13^

### Study Outcomes

We defined the primary outcome measure as the probability of moderate-to-severe Covid-19 hospitalization or Covid-19 death during the early Omicron wave, stratified by age group (12-59 years, 60-79 years, ≥80 years), immune status (severely immunocompromised, immunocompetent), and time since receipt of the last vaccine dose (0-3 months, 3-6 months, >6 months, and unvaccinated). For a comprehensive understanding of the total benefit of vaccination, we calculated for each subgroup the number of vaccinations required to prevent one severe Covid-19 hospitalization or death (NNV).

We computed the NNV for different scenarios depending on vaccination timing and Covid-19 prevalence levels. Specifically, we examined the following scenarios: vaccination at the beginning of a wave (i.e., high transmission settings, and 0-3 months since the last vaccine administration), vaccination every six months during high and low prevalence periods, and vaccination every six months with a late Covid-19 resurgence (namely, four to five months after last administration).

We repeated these analyses for immunocompromised individuals (inclusive of mild or moderate and severe immunocompromise) (Supplementary Appendix pp 5-6).

### Study design

For each day during the study period, t, we classified individuals as ‘severely immunocompromised’ dynamically, based on their daily status. Likewise, we tracked the immunity level achieved by vaccination on a daily basis within four categories: 0-3 months, 3-6 months and >6 months from the last vaccine dose uptake (either the second dose of the primary series or a booster dose), and unvaccinated (individuals who did not complete the primary series were considered unvaccinated). We determined the vaccination status seven days post inoculation, following studies suggesting that clinically relevant immunity is built during this window.^14,15^ Namely, we considered the vaccine dose to be effective only seven days after administration. We also stratified the population by age group (12-59 years, 60-79 years, and ≥80 years. Altogether, we classified individual days based on 24 (3 × 2 × 4) subgroups.

We then counted for each subgroup the total number of Covid-19 events (i.e., severe Covid-19 hospitalization or death) among the eligible populations on day t. To be eligible on day t, individuals had to be alive on day t-1, and not have had a Covid-19 hospitalization record between day 0 and day t-1. As previous Covid-19 illnesses protect against subsequent severe Covid-19 hospitalization or death, for each day t, we excluded individuals who tested positive for Covid-19 between the time ofreceiving the last vaccine dose and day t-30.^16,17^

### Statistical analysis

The risk of severe Covid-19 hospitalization, or death for each subgroup i, *p*_*i*_, is computed as the ratio between the number of total events and the accumulated number of eligible individuals throughout the examined period. The 95% confidence interval (Cl) was calculated assummg *β* distribution,

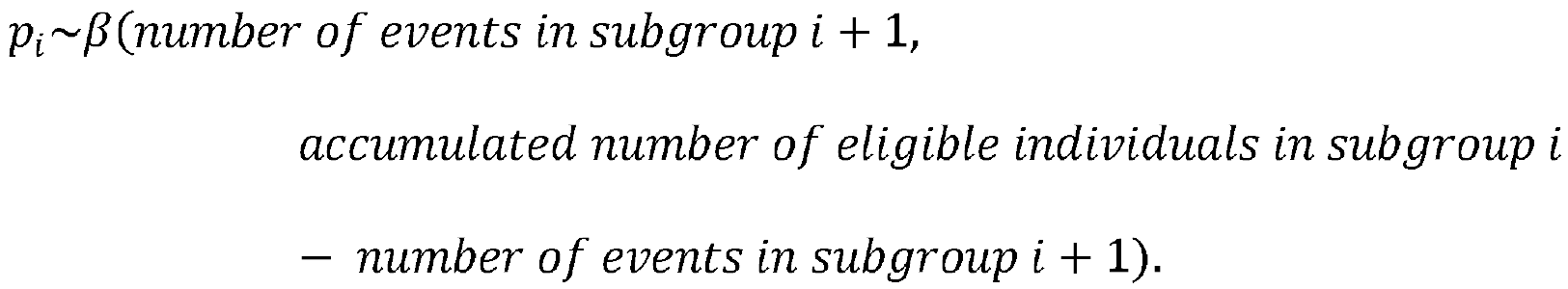

We computed the NNV for different scenarios depending on vaccination timing and Covid-19 transmission levels. For each scenario, the NNV was calculated as the inverse of the ARR (Absolute Risk Reduction). Specifically, for vaccinations administered at the beginning of a wave, the ARR was determined as the difference between the risk for individuals who received their last vaccination >6 months ago and the risk for individuals who were 0-3 months from their last vaccination. For vaccination every six months during high prevalence periods, the ARR was calculated as the difference between the risk for individuals who received their last vaccination >6 months ago and the risk for those vaccinated between 0-6 months ago. For the low prevalence periods, we multiplied the NNV of high prevalence settings by the ratio between the general number of severe Covid-19 hospitalizations during the early Omicron wave and the number of moderate or severe hospitalizations during low Covid-19 prevalence.

For the NNV, 95% CI were computed as follows: First, we drew 10,000 samples of percentile points. Then for each percentile point, we obtained the matching risk for each subgroup (assuming *β*) distribution and we calculated the relevant measure accordingly. Finally, the 95% CI was determined as the values between the 2.5^th^ percentile and the 97.5^th^ percentile.

## RESULTS

### Participant population

The cohort consisted of 3,381,480 CHS members, out of which 208,597 were severely immunocompromised and 3,172,883 were immunocompetent. Immunocompromised individuals were older than the immunocompetent with mean ages of 69 and 39, respectively. Likewise, immunocompromised individuals had more comorbidities compared to immunocompetent individuals, with mean number of comorbidities 2.09 and 0.71, respectively.

Within the severely immunocompromised cohort, females were overrepresented. Active malignancies were the most common immunocompromising conditions. Most participants were vaccinated in both immune status groups, though immunocompromised persons were more likely to have received second and third booster doses, per CDC guidelines. During the study period (early Omicron), 1 in 327 severely immunocompromised people died of Covid-19, compared to **1** in 1,953 immunocompetent people (Table 1).

**Table 1.** Cohort characteristics

| Characteristic | Severely immunocompromised* (n=208,597) | Immunocompetent** (n=3,172,883) |
| --- | --- | --- |
| **Sex** |  |  |
| Female | 58.7% (122,500) | 51.1% (1,620,207) |
| Male | 41.3% (86,097) | 48.9% (1,552,676) |
| **Age group** |  |  |
| Median (IQR) | 69 (56-77) | 39 (25-57) |
| 12-59 yr | 29.7% (61,920) | 78.0% (2,473,465) |
| 60-79 yr | 51.1% (106,499) | 17.8% (564,735) |
| 80+ yr | 19.3% (40,178) | 4.2% (134,683) |
| **Sociodemographic score\*\*\*** |  |  |
| 1-4 | 22.6% (47,167) | 36.3% (1,151,513) |
| 5-7 | 47.0% (97,967) | 40.6% (1,288,659) |
| 8-10 | 27.1% (56,601) | 18.2% (577,428) |
| Unspecified | 3.3% (6,862) | 4.9% (155,283) |
| **Comorbidity index&** |  |  |
| 0 | 27.9% (58,282) | 66.1% (2,097,884) |
| 1 | 21.7% (45,164) | 18.2% (578,919) |
| 2 | 16.8% (34,968) | 6.9% (219,392) |
| 3+ | 33.6% (70,183) | 8.7% (276,688) |
| **Population sector** |  |  |
| General Jewish | 84.6% (176,406) | 67.0% (2,124,792) |
| Ultra-Orthodox Jewish | 3.7% (7,683) | 5.5% (173,784) |
| Arab | 11.7% (24,508) | 27.6% (874,307) |
| **COVID-19 death** |  |  |
| Yes | 0.4% (858) | 0.1% (1,625) |
| No | 99.6% (207,739) | 99.9% (3,171,258) |
| **Immunocompromising condition** |  |  |
| Organ transplant | 4.6% (9,580) | 100.0% (3,172,883) |
| Hematologic malignancy | 89.3% (186,183) | 100.0% (3,172,883) |
| Active malignancy | 12.1% (25,222) | 100.0% (3,172,883) |
| **Vaccination status** |  |  |
| Unvaccinated | 9.7% (20,284) | 24.5% (776,886) |
| Primary series | 8.2% (17,138) | 18.4% (583,913) |
| First booster | 39.4% (82,087) | 45.5% (1,445,230) |
| Second booster | 24.2% (50,479) | 7.6% (241,459) |
| Third booster | 18.5% (38,609) | 4.0% (125,395) |

\* Severely immunocompromised includes people with an active malignancy, receipt of a solid-organ transplant, individuals diagnosed with hematopoietic malignancies (i.e., chronic lymphocytic leukemia, non-Hodgkin lymphoma, multiple myeloma, acute leukemia), or people diagnosed with cystic fibrosis or sickle cell anemia.
\*\* Immunocompetent includes individuals who are not severely immunocompromised or do not have the following conditions: Rheumatoid Arthritis, Ulcerative Colitis, Crohn's Disease, Celiac Disease, and Systemic Lupus Erythematosus.
\*\*\* Score for socioeconomic status (ranging from 1 [lowest] to 10 [highest]; details are provided in the Supplementary Appendix p. 2)
& Comorbidity index is the number of not immunosuppressive conditions: cardiovascular disease, diabetes, hypertension, chronic lung disease, chronic kidney disease and additional chronic diseases ( Supplementary Appendix pp 2-3)

### Absolute Risk for Severe Covid-19 Outcomes

The absolute risk of Covid-19-related moderate-to-severe hospitalization or death during the early Omicron wave increased with age, immune status (within each age group), and time since receipt of the last vaccine dose (observed at >3 months since receipt) (Figure 1). Unvaccinated people had the highest risk in all sub-groups.

**Figure 1.**
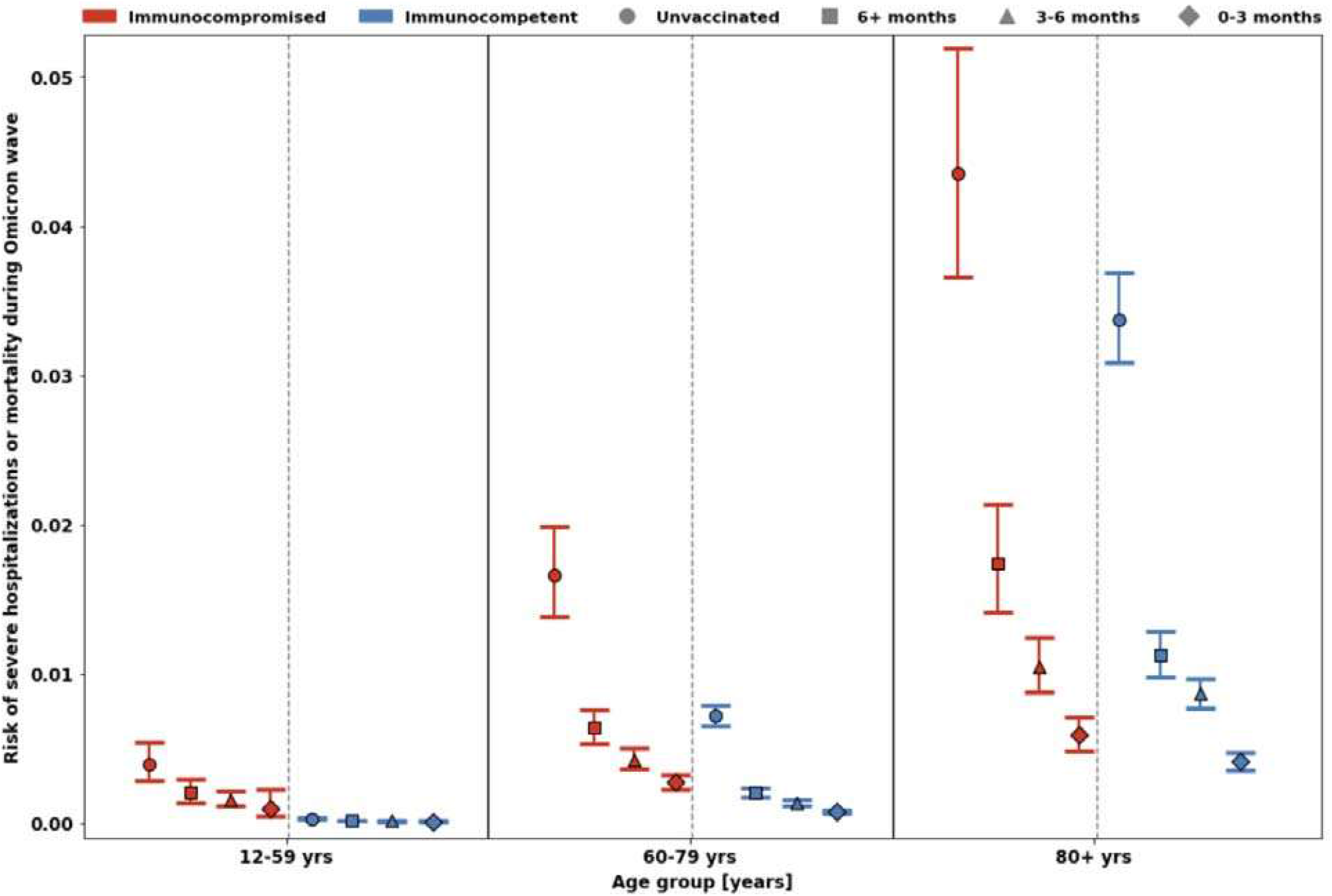
The risk of severe Covid-19-associated hospitalization or mortality is shown by age, immune status, and time since last vaccine dose. Immunocompromised groups are shown in red and immunocompetent groups are shown in blue. Unvaccinated persons are shown as circles, persons last vaccinated >6 months prior to the early Omicron wave are shown as boxes, persons last vaccinated 3-6 months prior to the early Omicron wave are shown as triangles, and persons last vaccinated 0-3 months prior to the early Omicron wave are shown as diamonds. Left: ages 12-59 years; Middle: ages 60-79 years. Right: ages ≥80 years. 95% confidence intervals are shown. A similar analysis for severely and mild or moderate immunocompromised individuals is shown in Figure S1.

Within each age group, immunocompromised persons had higher moderate-to-severe hospitalization and death risks, but that disparity increased with decreasing age. The excessive risk of immunocompromised subjects was 12.95-fold (95% CI 11.21-15.03) in the 12-59 age group, 3.15-fold (95% CI 2.99-3.33) in the 60-79 age group, and 1.55-fold (95% CI 1.44-1.66) in the >80 age group. When individuals with mild-to-moderate immunocompromise were added to the analysis, the absolute risks for severe Covid-19 outcomes were lower (Figure S1).

Younger immunocompromised people last vaccinated >6 months prior had the greatest disparity in outcomes (using immunocompetent persons ages ≥80 years last boosted >6 months prior as the reference standard). The rate of moderate-to-severe hospitalization or mortality during the initial Omicron wave among immunocompromised persons aged 12-59 (last dose >6 months prior) was 17.9% that of not recently boosted immunocompetent persons ages ≥80 (last dose >6 months prior) during that period. Meanwhile, among immunocompetent people ages 12-59 (last dose >6 months prior), the rate of moderate-to-severe hospitalization or mortality was only 1.4% that of the rate observed among immunocompetent persons ages ≥80 (last dose >6 months prior). Vaccinations narrowed (ages 12-59 and ages 60-79) or eliminated these disparities (ages ≥80) temporarily, but waning was apparent after 3 months.

### Number Needed to Vaccinate to Prevent Severe Covid-19 Events

The number of people needed to vaccinate (NNV) to prevent one severe Covid-19 hospitalization or death (6-month cadence) varied greatly by age and immune status and was contingent on various disease prevalence scenarios. Among the severely immunocompromised, doses administered at the start of the Omicron wave had an NNV ranging from 87 (95% CI 70-109) in persons ages ≥80 to 1,037 (95% CI 999-1,513) in persons ages 12-59. In the three lower prevalence scenarios (Table 2), the NNV for 6-month booster cadencing remained favorable for immunocompromised people in all age groups and for immunocompetent people ages ≥60. However, NNVs for immunocompetent people <age 60 were extremely high 98,934 (95% CI 98,188-104,814) during low prevalence periods. When individuals with mild-to-moderate immunocompromise were included in the analysis, the NNVs were much higher in certain scenarios (Table Sl). NNVs for mortality alone are also shown (Table 2, Table Sl).

**Table 2.**
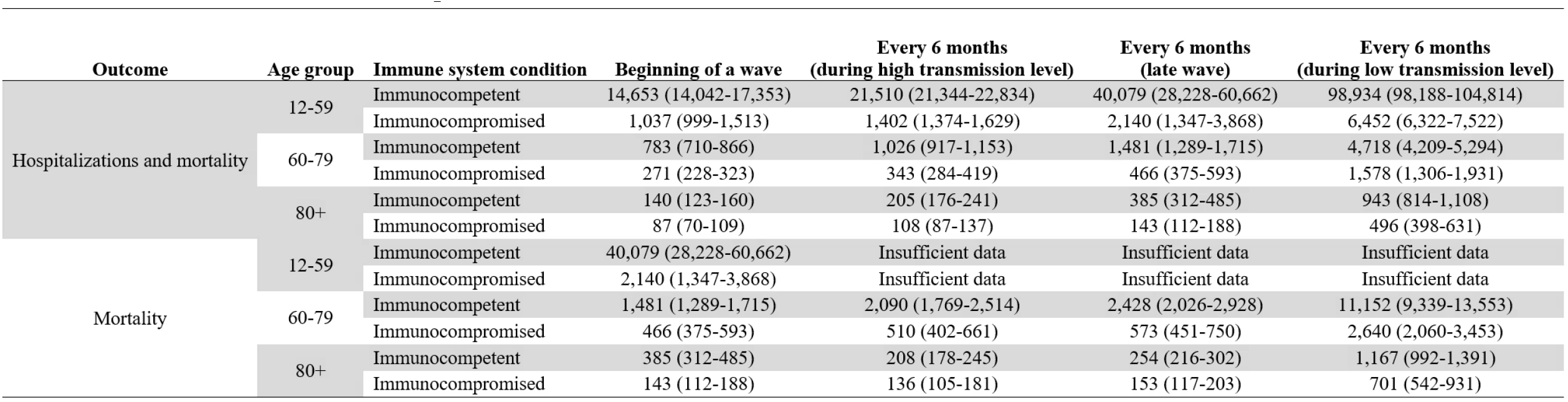
Number needed to vaccinate to prevent a severe COVID-19 event.

## DISCUSSION

The present analysis indicates that a routine 6-month cadence for Covid-19 booster vaccinations for the highest-risk groups may be warranted, even during times of relatively lower Covid-19 prevalence. Moreover, the NNV analysis strongly implies that a 2^nd^ bivalent booster dose is currently warranted among persons with severe immune compromise and advanced age who are >6 months from their first bivalent dose (and possibly >3 months, depending on immune status and prevalence).

These findings highlight the high absolute risk that severely immunocompromised people have compared to immunocompetent people. For example, severely immunocompromised persons ages 12-59 last vaccinated >6 months before the Omicron wave had approximately 48.9% the risk as that ofrecently vaccinated immunocompetent persons ages ≥80 at that time (Figure 1, compare the red square in the far-left panel to blue diamond in the far-right panel). However, short-term effectiveness was adequate among immunocompromised people such that booster doses substantially closed the disparity gap to same-aged immunocompetent persons (ages 12-59 and ages 60-79), or completely eliminated the gap (ages ≥80), albeit temporarily.

We found that the risk disparity between the immunocompromised and immunocompetent decreased dramatically with age, likely because baseline risks among the immunocompetent population increases dramatically with age (i.e., the baseline risk of the immunocompetent is very low among persons ages 12-59, and higher among 60-79 and ≥80-year-old populations).

Additionally, our results demonstrated that immunocompromised individuals who are unvaccinated and unvaccinated individuals ages ≥60 years have the highest risk for severe Covid-19 outcomes. We found that unvaccinated immunocompromised persons ages ≥80 years or older had a risk of 4.4% (95% CI: 3.7%-2.2%) for moderate-to-severe Covid-19 hospitalization or death during the early Omicron period (Figure 1, right).

### Comparison to Current Evidence

Immunocompromised individuals are at substantially higher risk of serious Covid-19 outcomes compared to the immunocompetent population.^18^ In a large surveillance network in the United States during the Omicron period, persons with moderate-to-severe immune compromise accounted for 12.2% of Covid-19-associated hospitalizations, despite historical data suggesting that this group represents only 2.7% of the general population. ^19,20^

The effectiveness of bivalent vaccines in preventing severe Covid-19 outcomes among the general population age >65 years has been demonstrated in Israel, but the calculated NNVs determined in that study reflected pooled risks between all older persons, regardless of advanced age (e.g., >80 years) or immune status.^21^

While the effectiveness of the bivalent booster now in use may be marginally higher (and the waning somewhat slower) compared to the previously in-use monovalent booster, the overall epidemiologic difference (monovalent versus bivalent) currently appears minimal. In addition, while boosters are effective against Covid-19-associated hospitalization, the effect size of the additional benefit (in comparison to vaccinated but not boosted persons) appears relatively small and short-lived among the general older population.^22,23^ Nevertheless, when considering the absolute risks and waning booster effectiveness, the marginal improvement offered by bivalent vaccines do not appear to have altered the conclusions indicated by the present analysis.

### Implications for Policy

Our study establishes the population benefit and likely necessity of a 6-month booster cadence (at least) for persons with severe immunocompromise and immunocompetent older populations ages >60 years. Given that recent evidence demonstrates that severe adverse events following administration of booster vaccine doses are extremely rare in the high-risk population, a 6-month booster cadence is reasonable compared to the high burdens of Covid-19.^27,28^

The utility that additional boosting among the highest risk groups may provide (i.e., boosting every 3-4 months, for example) remains unexplored. However, our data show that a dramatic drop in the protection added by boosters occurred after just 0-3 months. This implies the need for a further assessment ofNNVs for even shorter booster cadences for the highest-risk groups studied here. Given that the difference in NNVs are contingent on prevalence, a twice or three-times per-year booster cadence for the highest-risk groups further augmented by ‘just-in time” wave-triggered booster campaigns may be warranted.

The concept of waiting to administer additional boosters until a large wave occurs has been discussed by policy experts but is controversial. However, given that millions of vaccine doses per day were administered in the United States during the initial vaccine rollout, it is feasible that a high proportion of the highest-risk population could be boosted quickly, if ground conditions warranted.

We propose that once a 6-month booster cadence (at a minimum) for the highest risk groups is in place, the safety, effectiveness, and reasonableness of further boosting (e.g., routine cadence or ‘just-in-time” doses offered during waves occurring >3 months after the last receipt of a booster) should be explored for the highest-risk population. Acknowledged risks of such an endeavor would include the possibility of negative immune imprinting and new adverse events, albeit these are less likely to occur in severely immunocompromised and older persons, precisely because vaccines are less immunogenic in this population.^29,30^

While the absolute risk reduction among the younger severely immunocompromised population was lower than that of older severely immunocompromised people, a “years of lost” life analysis would likely largely or completely negate any differences. Therefore, for the purposes of prioritization, it is reasonable to make one policy for severely immunocompromised people, regardless of age. Conversely, the baseline risks and booster-associated absolute risk reductions among immunocompetent groups are divergent enough that age-specific guidance remain worthwhile. In addition, the effect size of boosting persons with mild immunocompromise was smaller (Table S1), indicating people with mild-to-moderate immunocompromise may need to be considered separately.

Currently, in the United Kingdom and Canada, second bivalent booster doses are being offered to similarly high-risk groups as the study population assessed here. The data here support these policies and argue for expanding them in other nations with similar demographic risks.^31,32^

### Limitations

Our study has several limitations. First, these data assessed outcomes during periods in which the dominant circulating variants were Omicron, while available booster vaccines were monovalent to the receptor-binding domain of the Wuhan strain only. However, as above, recent data on the bivalent vaccine do not appear to have markedly changed the landscape.^21,23,24^ Second, we did not assess in-group differences by traditional Covid-19 risk factors.^25^ Future studies are warranted to identify a comprehensive highest-risk cohort that might benefit from a 6-month booster cadence or wave-triggered boosts, and those (including persons above age 60) who may not. Third, asymptomatic infections are not completely reported (and could not be explicitly accounted for in our analysis). This may be relevant, as previous infection provides further protection against severe Covid-19 outcomes. However, if we had been able to exclude asymptomatic cases (focusing only on the population that remained uninfected after inoculation), the risks and effectiveness described here would be anticipated to be even higher, strengthening the conclusions the present results imply. Furthermore, we did not account for the receipt of effective antiviral treatments against severe Covid-19, Even during the vaccine era.^26^ Finally, our data reflect the Israeli population which may differ in characteristic compared with other nations. For example, in the US, immunocompromised individuals tend to be younger compared to our individuals included in our dataset.^20^

## Conclusions

Our study provides evidence for the potential benefit of a routine 6-month cadence for Covid-19 boosters (and possibly more frequently) for the highest-risk groups, even during relatively lower Covid-19 prevalence. Our findings are derived from the disproportional risk of severe Covid-19 outcomes to the immunocompromised populations and the potentially high, albeit waning, vaccine benefit.

## Data Availability

All data produced in the present study are available upon reasonable request to the authors to the extent permitted by law/privacy agreements.

## Notes

### Competing Interest Statement

The authors have declared no competing interest.

### Funding Statement

This study did not receive any funding

### Author Declarations

The IRB of Clalit Health Services gave ethical approval for this work.

## References

1. Tenforde MW. Effectiveness of a Third Dose of Pfizer-BioNTech and Modema Vaccines in Preventing COVID-19 Hospitalization Among Immunocompetent and Immunocompromised Adults-United States, August-December 2021. MMWR Morb Mortal Wkly Rep. 2022;71. doi:10.15585/mmwr.mm7104a2

2. Ferdinands JM, Rao S, Dixon BE, et al. Waning of vaccine effectiveness against moderate and severe covid-19 among adults in the US from the VISION network: test negative, case control study. BMJ. 2022;379:e072141. doi:10.1136/bmj-2022-072141

3. Britton A. Effectiveness of COVID-19 mRNA Vaccines Against COVID-19-Associated Hospitalizations Among Immunocompromised Adults During SARS-CoV-2 Omicron Predominance - VISION Network, 10 States, December 2021-August 2022. MMWR Morb Mortal Wkly Rep. 2022;71. doi:10.15585/mmwr.mm7142a4

4. Research C for DE and. FDA announces Evusheld is not currently authorized for emergency use in the U.S. FDA. Published online January 25, 2023. Accessed April 14, 2023. https://www.fda.gov/drugs/drug-safety-and-availability/fda-announces-evusheld-notcurrently-authorized-emergency-use-us

5. Commissioner O of the. Coronavirus (COVID-19) Update: FDA Authorizes Additional Vaccine Dose for Certain Immunocompromised Individuals. FDA. Published August 16, 2021. Accessed April 7, 2023. https://www.fda.gov/news-events/press-announcements/coronavirus-covid-19-update-fda-authorizes-additional-vaccine-dosecertain-immunocompromised

6. CDC/ACIP. ACIP COVID-19 Vaccine Recommendations I CDC. Published December 27, 2022. Accessed April 7, 2023. https://www.cdc.gov/vaccines/hcp/acip-recs/vaccspecific/covid-19.html

7. CDC. COVID19-vaccination-schedule-immunocompromised. Accessed April 7, 2023. https://www.cdc.gov/vaccines/covid-19/images/COVID19-vaccination-scheduleimmunocompromised.png

8. CDC. COVID-19 Vaccination: Clinical & Professional Resources. Centers for Disease Control and Prevention. Published May 13, 2022. Accessed April 7, 2023. https://www.cdc.gov/vaccines/covid-19/index.html

9. Oliver S. Considerations for Future Planning.

10. Cohen R, Rabin H. Membership in a health fund, 2016. http://www.btl.gov.il. Published August 2017. Accessed April 14, 2023. http://www.btl.gov.il:80/Publications/survey/Pages/seker_289.aspx

11. CDC. ACIP Altered Immunocompetence Guidelines for Immunizations I CDC. Published April 12, 2023. Accessed April 14, 2023. https://www.cdc.gov/vaccines/hcp/aciprecs/general-recs/immunocompetence.html

12. Our World in Data. SARS-CoV-2 sequences by variant. Our World in Data. Accessed April 14, 2023. https://ourworldindata.org/grapher/covid-variants-bar

13. Israeli Ministry of Health. COVID-19 Data Tracker. Accessed April 14, 2023. https://datadashboard.health.gov.il/COVID-19/general

14. Arbel R, Sergienko R, Friger M, et al. Effectiveness of a second BNTl 62b2 booster vaccine against hospitalization and death from COVID-19 in adults aged over 60 years. Nat Med. 2022;28(7):1486–1490. doi:10.1038/s41591-022-01832-0

15. Grewal R, Kitchen SA, Nguyen L, et al. Effectiveness of a fourth dose of covid-19 mRNA vaccine against the omicron variant among long term care residents in Ontario, Canada: test negative design study. BMJ. 2022;378:e071502. doi:10.1136/bmj-2022-071502

16. Tenforde MW, Link-Gelles R, Patel MM. Long-term Protection Associated With COVID-19 Vaccination and Prior Infection. JAMA. 2022;328(14):1402–1404. doi:10.1001/jama.2022.14660

17. Stein C, Nassereldine H, Sorensen RID, et al. Past SARS-CoV-2 infection protection against re-infection: a systematic review and meta-analysis. The Lancet. 2023;401(10379):833–842. doi:10.1016/SO140-6736(22)02465-5

18. Nevejan L, Ombelet S, Laenen L, et al. Severity of COVID-19 among Hospitalized Patients: Omicron Remains a Severe Threat for Immunocompromised Hosts. Viruses. 2022;14(12):2736. doi:10.3390/vl4122736

19. Singson JRC. Factors Associated with Severe Outcomes Among Immunocompromised Adults Hospitalized for COVID-19-COVID-NET, 10 States, March 2020-February 2022. MMWR Morb Mortal Wkly Rep. 2022;71. doi:10.15585/mmwr.mm7127a3

20. Harpaz R, Dahl RM, Dooling KL. Prevalence of Immunosuppression Among US Adults, 2013. JAMA. 2016;316(23):2547–2548. doi:10.1001/jama.2016.16477

21. Arbel R, Peretz A, Sergienko R, et al. Effectiveness of a bivalent mRNA vaccine booster dose to prevent severe COVID-19 outcomes: a retrospective cohort study. Lancet Infect Dis. 2023;0(0). doi:10.1016/S1473-3099(23)00122-6

22. Arbel R, Peretz A, Sergienko R, et al. Effectiveness of the Bivalent mRNA Vaccine in Preventing Severe COVID-19 Outcomes: An Observational Cohort Study. SSRN Electron J. Published online 2022. doi:10.2139/ssm.4314067

23. Lin DY, Xu Y, Gu Y, et al. Effectiveness of Bivalent Boosters against Severe Omicron Infection. N Engl J Med. 2023;388(8):764–766. doi:10.1056/NEJMc2215471

24. Lin DY, Xu Y, Gu Y, Zeng D, Sunny SK, Moore Z. Durability of Bivalent Boosters against Omicron Subvariants. N Engl J Med. 2023;0(0):null. doi:10.1056/NEJMc2302462

25. Williamson EJ, Walker AJ, Bhaskaran K, et al. Factors associated with COVID-19-related death using OpenSAFELY. Nature. 2020;584(7821):430–436. doi:10.1038/s41586-020-2521-4

26. Arbel R, Wolff Sagy Y, Hoshen M, et al. Nirmatrelvir Use and Severe Covid-19 Outcomes during the Omicron Surge. N Engl J Med. 2022;387(9):790–798. doi:10.1056/NEJMoa2204919

27. Yamin D, Yechezkel M, Arbel R, et al. Safety of COVID-19 Monovalent and Bivalent BNT162b2 mRNA Vaccine Boosters for Adults 60 Years and Above: A Large-Scale Retrospective Study. Published online January 25, 2023. doi:10.2139/ssm.4336133

28. Yechezkel M, Mofaz M, Painsky A, et al. Safety of the fourth COVID-19 BNT162b2 mRNA (second booster) vaccine: a prospective and retrospective cohort study. Lancet Respir Med. 2023;11(2):139–150. doi:l 0.1016/S2213-2600(22)00407-6

29. Chemaitelly H, Ayoub HH, Tang P, et al. Long-term COVID-19 booster effectiveness by infection history and clinical vulnerability and immune imprinting: a retrospective population-based cohort study. Lancet Infect Dis. 2023;0(0). doi:10.1016/S1473-3099(23)00058-0

30. Hause AM. Safety Monitoring of COVID-19 mRNA Vaccine First Booster Doses Among Persons Aged 12 Years with Presumed Immunocompromise Status - United States, January 12, 2022-March 28, 2022. MMWR Morb Mortal Wkly Rep. 2022;71. doi:10.15585/mmwr.mm7128a3

31. Canada PHA of. COVID-19 vaccine: Canadian Immunization Guide. Published December 23, 2021. Accessed April 7, 2023. https://www.canada.ca/en/publichealth/services/publications/healthy-living/canadian-immunization-guide-part-4-activevaccines/page-26-covid-l9-vaccine.html

32. GOV.UK. A guide to the COVID-19 spring booster 2023. Accessed April 7, 2023. https://www.gov.uk/government/publications/covid-19-vaccination-spring-boosterresources/a-guide-to-the-covid-l9-spring-booster-2023

