## Supplemental Appendix for "Covid-19 Vaccine Booster Cadence by Immunocompromised Status"

### Appendix A – Study protocol

***Description of the data***

Data will be extracted from the Clalit Healthcare Services (CHS) database. CHS is the largest nationwide health plan (payer-provider) representing a half of the population in Israel. The CHS database contains longitudinal data on a stable population of 4·7 million people since 1993 (with <1%/year moving out) ^1^. Data are automatically collected and includes comprehensive laboratory data from a single central lab, full pharmacy prescription and purchase data, and extensive demographic data on each patient. CHS uses the International Classification of Diseases, Ninth Revision, Clinical Modification (ICD-9-CM) coding systems as well as self-developed coding systems to provide more granular diagnostic information beyond the ICD codes. Medications are coded according to the Israeli coding system with translations to anatomical therapeutic chemical (ATC) classification system wherever available. Procedures are coded using Current Procedural Terminology (CPT) codes. We will access to the following data for each patient:

- Socio-demographics
- Sex (binary)
- Age (year of birth)
- Socioeconomic status by address and according clinic when address is missing) (scale 1-10)
- Sector (clinic level data - Arab / Jewish/ Ultra-Orthodox Jewish)
- Nursing home residency (binary)
- Comorbidities
- Chronic diseases (binary classifcation)
- History of malignancies and active malignancies
- Dementia
- Cardiovascular diseases (ischaemic heart disease, cerebrovascular disease/ all cardio sub registries).
- Diabetes (taken from CRI registry)
- Hypertension (taken from CRI registry)
- Asthma
- Chronic Lung Disease
- Rheumatologic diseases
- Chronic Kidney Disease
- Immunocompromised Status
- Chronic Liver Disease
- Organ transplantation
- Additional Chronic Diseases
- Vaccination records
- Laboratory test results ( binary classification for existence of infectious diseases)
- Hospitalization history ({ admission data, primary service, duration, ICD diagnosis code })
- Outpatient history (admission data, primary service, ICD diagnosis code)
- BMI ({date ,value}}
- Smoking status {date, yes/no}

*Explanation on the Socio-Economic Status (SES)*

Measure SES was based on the small statistical areas (SSA) used in the 2008 Israeli census ^2^. The SSAs contain 3000–4000 people and are created to maintain homogeneity in terms of the sociodemographic composition of the population. The Israeli Central Bureau of Statistics (CBS) utilized demography, education, employment, housing conditions, and income to define the SSAs, and these were grouped into 20 categories ^3^. This data was updated by the POINTS Location Intelligence Company to improve the accuracy of the SES measure, using up-to date sociodemographic, commercial, and housing data. The entire CHS population was grouped into ten categories, ranging from 1 (lowest) to 10 (highest).

***Data collection and storage***

We will receive access to the data from the medical records of all random members of Clalit Healthcare Services aged 12 years and older. CHS is the largest nationwide health plan (payer-provider) representing a half of the population in Israel. The CHS database contains longitudinal data on a stable population of 4·7 million people since 1993 (with <1%/year moving out). Data are automatically collected and includes comprehensive laboratory data from a single central lab, full pharmacy prescription and purchase data, and extensive demographic data on each patient. CHS uses the International Classification of Diseases, Ninth Revision, Clinical Modification (ICD-9-CM) coding systems as well as self-developed coding systems to provide more granular diagnostic information beyond the ICD codes. Medications are coded according to the Israeli coding system with translations to anatomical therapeutic chemical (ATC) classification system wherever available. Procedures are coded using Current Procedural Terminology (CPT) codes. As for the medical data, we will receive access to the EMR data after the following pseudonymisation procedures:

1. Healthcare identification number of the members will be coded.
2. Only year of birth is provided.
3. Free text is removed. This means any text that was types/recorded/scanned manually by healthcare staff, and is not structured in the electronic system. This includes any documented conversations between healthcare staff and patient or summary of from meetings.
4. No audio, photos including scanned text, or video contents are provided.

The data access will be conducted at the CHS. The data are coded, viewed, stored and process only within the Clalit research room. The researchers will connect to the research room via Horizon client platform, which is approved by the Ministry of Health. The user connects through a secure connection using Lightweight Directory Access Protocol and two factor authentication system.

***Outcomes***

We define the primary outcome measures as the probability of exhibiting severe COVID-19 hospitalizations, or COVID-19 deaths during the early omicron wave stratified by age group (12-59 years, 60-79 years, and ≥80 years), immune status (immunocompromised, immunocompetent), and time since receipt of the last vaccine dose (0-3 months, 3-6 months, >6 months, and unvaccinated), which lasted between December 20,2021 and April 5, 2022. For a comprehensive understanding of the total benefit of vaccination, we will calculate for each subgroup the number of vaccines that are required to prevent one case of severe COVID-19 hospitalization or death (NNV) for those.

We will compute the NNV for different scenarios depending on vaccination timing and COVID-19 transmission levels. Specifically, we will examine the following scenarios: vaccination at the beginning of a wave (i.e., high transmission settings, and 0-3 months passed since the last vaccine administration), vaccination every six months during high and low transmission levels, and vaccination every six months with a late COVID-19 resurgence (namely, four to five months after administration).

### Appendix B – Study Design

To be included in our analysis, individuals need to be active CHS members during the entire study period, except for death. Additionally, individuals have to be unvaccinated or vaccinated with at least one dose of the BNT162b2 mRNA COVID-19 vaccine. We will exclude individuals who received any other COVID-19 vaccine dose than the BNT162b2 mRNA COVID-19 vaccine.

For each day during the study period, t, we will classify individuals as ‘severely immunocompromised’ in a dynamic way based on their daily status. The definition of ‘severely immunocompromised’ is based on the US CDC’s and the Advisory Committee on Immunization Practices’ (ACIP) definition ^4,5^. Specifically, severely immunocompromised individuals include people with an active malignancy, receipt of a solid-organ transplant, individuals diagnosed with hematopoietic malignancies (i.e., chronic lymphocytic leukemia, non-Hodgkin lymphoma, multiple myeloma, acute leukemia), or people diagnosed with cystic fibrosis or sickle cell anemia. We will classify individuals as immunocompetent if they are not immunocompromised (mild or moderate or severely). Mild or moderate immunocompromised individuals include people with at least one of the following conditions: Rheumatoid Arthritis, Ulcerative Colitis, Crohn’s Disease, Celiac Disease, and Systemic Lupus Erythematosus. This definition is based on the on the US CDC’s and the Advisory Committee on Immunization Practices’ (ACIP) definition ^4,5^. Likewise, we will track on a daily basis the immunity level that may be achieved by vaccination based on the four categories: 0-3 months, 3-6 months and >6 months from the last vaccine dose uptake (second dose of the primary series or booster dose), and unvaccinated. We will count the vaccination status seven days post inoculation following studies that suggest immunity is built following at least seven days after vaccination ^6,7^. Namely, we considered the vaccine dose to be effective only seven days after the administration. We will also stratify the population by age group (12-59 years, 60-79 years, and $\geq80$ years). Altogether, we will classify individual days based on 24 ($3\times2\times4$) subgroups.

We then will count for each subgroup the total number of COVID-19 events (i.e., severe COVID-19 hospitalization or death) among the eligible populations on day t. To be eligible on day t, individuals must be alive at day t-1, and not have a COVID-19 hospitalization record between day 0 and day t-1. Previous COVID-19 infections may protect from severe COVID-19 hospitalization or death ^8,9^. Thus, for each day t, we will exclud individuals who tested positive for COVID-19 between the time of receiving the last vaccine dose and day t-30.

### Appendix C - Results for all immunocompromised individuals (severely and mild or moderate) aged 12 years and above

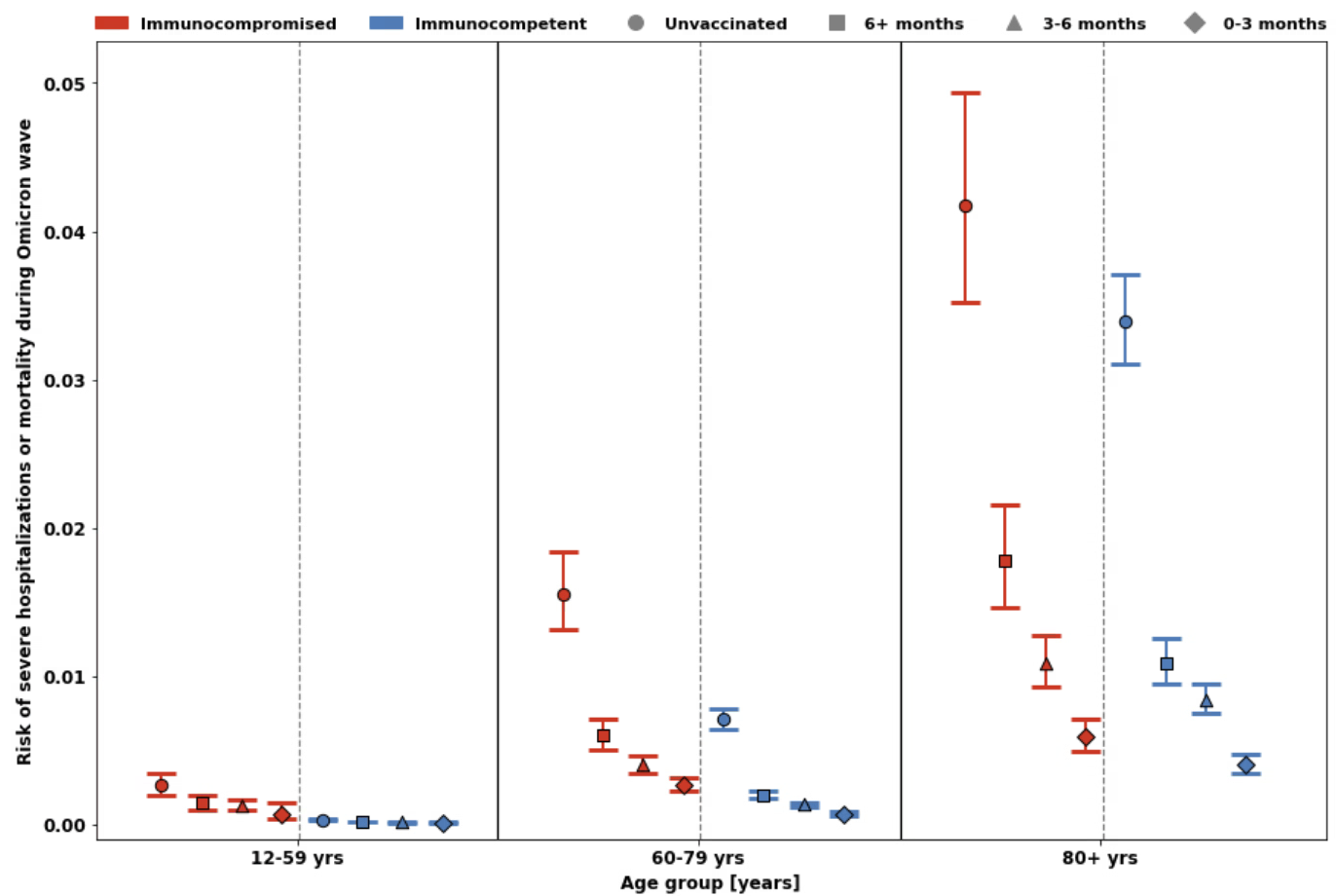

Figure S1. The risk of severe Covid-19-associated hospitalization or mortality is shown by age, immune status, and time since last vaccine dose. Immunocompromised groups are shown in red and immunocompetent groups are shown in blue. Unvaccinated groups are shown as circles, those last vaccinated >6 months prior to the early Omicron wave are shown as boxes, those last vaccinated 3-6 months prior to the early Omicron wave are shown as triangles, and those last vaccinated 0-3 months prior to the early Omicron wave are shown as diamonds. Left: ages 12-59 years; Middle: ages 60-79 years. Right: ages ≥80 years. 95% confidence intervals are shown.

**Table S1**. Number needed to vaccinate to prevent a severe COVID-19 event among severely and mild or moderate immunocompromised, and immunocompetent individuals

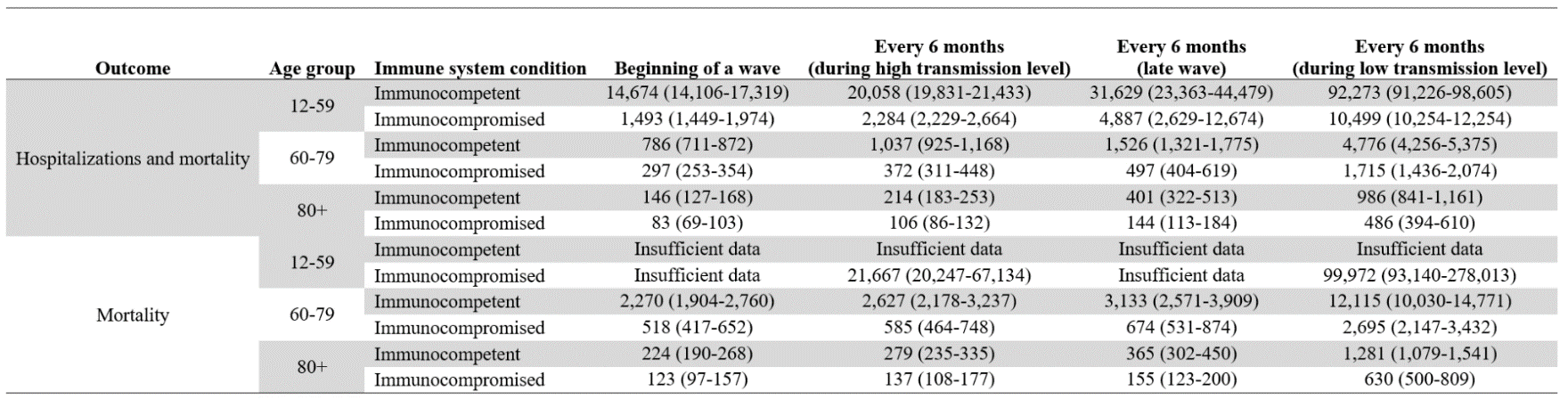

### References

1.	Cohen R, Rabin H. National Insurance Institute of Israel. Membership in sick funds 2016. August 2017. (In Hebrew) [Internet]. [cited 2023 Jan 16];Available from: https://www.btl.gov.il/Publications/survey/Documents/seker289/seker_289.pdf

2.	Arbel R, Hammerman A, Sergienko R, et al. BNT162b2 Vaccine Booster and Mortality Due to Covid-19. New England Journal of Medicine [Internet] 2021 [cited 2023 Mar 6];385(26):2413–20. Available from: https://www.nejm.org/doi/full/10.1056/NEJMoa2115624

3.	CHARACTERIZATION AND CLASSIFICATION OF GEOGRAPHICAL UNITS BY THE SOCIO-ECONOMIC LEVEL OF THE POPULATION 2008 [Internet]. [cited 2023 Mar 6];Available from: https://www.cbs.gov.il/en/publications/Pages/2013/CHARACTERIZATION-AND%C2%A0CLASSIFICATION-OF%C2%A0GEOGRAPHICAL-UNITS%C2%A0BY-THE-SOCIO-ECONOMIC-LEVEL-OF-THE-POPULATION-2008.aspx

4.	People Who Are Immunocompromised | CDC [Internet]. [cited 2023 Apr 14];Available from: https://www.cdc.gov/coronavirus/2019-ncov/need-extra-precautions/people-who-are-immunocompromised.html

5.	Rubin LG, Levin MJ, Ljungman P, et al. 2013 IDSA clinical practice guideline for vaccination of the immunocompromised host. Clinical Infectious Diseases 2014;58(3).

6.	Grewal R, Kitchen SA, Nguyen L, et al. Effectiveness of a fourth dose of covid-19 mRNA vaccine against the omicron variant among long term care residents in Ontario, Canada: test negative design study. BMJ [Internet] 2022 [cited 2023 Apr 14];378. Available from: https://www.bmj.com/content/378/bmj-2022-071502

7.	Arbel R, Sergienko R, Friger M, et al. Effectiveness of a second BNT162b2 booster vaccine against hospitalization and death from COVID-19 in adults aged over 60 years. Nature Medicine 2022 28:7 [Internet] 2022 [cited 2023 Apr 14];28(7):1486–90. Available from: https://www.nature.com/articles/s41591-022-01832-0

8.	Tenforde MW, Link-Gelles R, Patel MM. Long-term Protection Associated With COVID-19 Vaccination and Prior Infection. JAMA [Internet] 2022 [cited 2023 Apr 14];328(14):1402–4. Available from: https://jamanetwork.com/journals/jama/fullarticle/2796894

9.	Stein C, Nassereldine H, Sorensen RJD, et al. Past SARS-CoV-2 infection protection against re-infection: a systematic review and meta-analysis. The Lancet [Internet] 2023 [cited 2023 Apr 14];401(10379):833–42. Available from: http://www.thelancet.com/article/S0140673622024655/fulltext
